# Sexually transmitted infection test positivity and associated factors in individuals tested at anonymous clinics in North Rhine-Westphalia state, Germany, 2021

**DOI:** 10.1101/2025.06.18.25329860

**Authors:** Katja Siling, Pia Grotegut, Emma M. Harding-Esch, Annette Jurke

**Affiliations:** Landeszentrum Gesundheit, NRW Centre for Health, Bochum, Germany; London School of Hygiene & Tropical Medicine (LSHTM), London, UK

## Abstract

Sexually transmitted infections (STIs) are increasing globally. To improve understanding of the epidemiology of STIs in key populations, we analysed sentinel surveillance data on STI test positivity among individuals tested using subsidised tests at anonymous walk-in clinics in North Rhine-Westphalia (NRW), Germany.

This was a cross-sectional analysis of sentinel surveillance data from all 53 local health authority anonymous walk-in clinics in NRW during 2021. Test positivity for HIV, syphilis, chlamydia, and gonorrhoea was analysed by key population group. We used univariable and multivariable logistic regression to examine factors associated with each infection.

Among 11,560 consultations, overall STI test positivity was 6.6%. Chlamydia was the most frequently detected (6.4%), followed by gonorrhoea (2.7%), syphilis (0.9%), and HIV (0.6%). We observed disparities in STI test positivity among key population groups. Men-who-have-sex-with-men (MSM) (aOR 1.93; 95%CI 1.38-2.68), sex work (aOR 2.77; 95%CI 1.9-3.96), and having an STI-positive partner (aOR 1.71; 95%CI 1.30-2.22) were associated with increased STI risk. Coinfections were rare (0.4%) and predominantly occurred among MSM.

Anonymous walk-in clinics reach diverse high-risk populations with substantial STI burdens. Findings support targeted prevention strategies for key populations and highlight the importance of comprehensive STI screening in anonymous settings for surveillance and case detection.

## Background

The global incidence of sexually transmitted infections (STIs) is increasing worldwide ^1, 2^. In Europe, *Chlamydia trachomatis* (CT) and *Neisseria gonorrhoeae* (NG) are the most commonly reported STIs ^3^. However, CT and NG data are not always routinely reported for national surveillance purposes. In Germany, antimicrobial-resistant NG has been notifiable since March 2020 ^4^, while CT and NG became notifiable in September 2022 ^5, 6^. However, as of March 2025, the system for reporting CT and NG diagnoses was not yet operational ^7^. Thus, most prevalence and incidence estimates are based on research studies. CT prevalence estimates for women and men attending gynaecological and urological clinics are low (0.59% and 0.36%, respectively) ^8, 9^. Higher CT prevalence is reported from clinic settings, around 5% in women and 3% in heterosexual men ^10, 11^. For NG, the reported prevalences are 1.5% in women ^12^ and 0.12% in men ^9^. In key populations, studies have focused primarily on men who have sex with men (MSM), reporting CT prevalences of 1.5-10.0% ^10, 11, 13, 14^ and NG prevalences of 1.9-8.9% ^11, 13, 14^. Prevalence of CT and NG in sex workers is estimated to be 6.8% and 3.2%, respectively ^15^, while peer-reviewed reports for prevalence in people who inject drugs (PWID) in Germany are lacking.

In contrast, HIV and syphilis are notifiable diseases in all European countries. German national surveillance data report the incidence of HIV and syphilis, but without the number of tests performed, which means that prevalence data are lacking. In 2023, a total of 3,321 new HIV infections were reported ^16^. The three most commonly reported likely routes of infection were sexual contact between men (MSM; 36.1%), heterosexual exposure abroad (17.9%), and injection drug use (5.9%). Of the 8,305 syphilis cases reported in 2022 ^17^, transmission among MSM was the most commonly reported likely route of infection (85.6%).

The availability and type of STI testing services in Germany vary between federal states and regions. Free screening for CT is offered to sexually active women under 25 years of age, and for HIV and syphilis to pregnant women, usually in their first trimester. STIs are primarily diagnosed and treated by clinicians, some of whom are organised into STI specialist centres. Health insurance usually covers the cost of a chlamydia test only if there are symptoms or a confirmed infection in a sexual partner. Comprehensive STI screening options for asymptomatic individuals without high-risk exposure are limited and usually not covered by statutory health insurance ^11^. Under the Infection Protection Act ^5^, local health authorities (LHA) are legally obliged to offer free counselling and testing for STIs to high-risk populations through anonymous walk-in clinics. These clinics primarily target people without health insurance but also serve vulnerable populations who face barriers to accessing healthcare services ^18^. In addition, some clinics offer STI testing to a wider population, with costs varying according to the funding structure.

Germany’s national STI strategy ^19^ aims to improve prevention, testing, and treatment of STIs by enhancing surveillance and addressing gaps in high-risk populations. The strategy emphasises the importance of strengthening data collection systems and expanding access to testing services, particularly for key populations who face disproportionate STI burdens. However, significant surveillance gaps persist, limiting a comprehensive understanding of STI epidemiology.

In North Rhine-Westphalia state (NRW), Germany’s most populous state, HIV and syphilis trends reflect overall national trends ^16, 17^. In 2021, all 53 anonymous walk-in clinics in NRW began offering state-funded CT and NG testing for the first time, in addition to existing free HIV and syphilis testing services. This expansion provided a unique opportunity to examine STI epidemiology across key populations accessing anonymous testing services. To describe the epidemiology of STIs and the populations reached by anonymous walk-in clinics in NRW, we analysed test positivity and factors associated with HIV, syphilis, CT and/or NG infections in individuals tested at these clinics.

## Methods

### Study design and population

This was a retrospective cross-sectional analysis of routinely collected sentinel surveillance data at all 53 LHA anonymous walk-in clinics in NRW between 1^st^ January 2021 – 31^st^ December 2021. We analysed anonymised data of individuals who were tested with free state-funded tests for HIV, syphilis, CT and/or NG.

### Data collection

During consultations, healthcare professionals filled in a paper questionnaire collecting socio-demographic data, including the individual’s age, sex, country of origin, high-risk exposure (i.e., MSM, heterosexual contact, sex work, injection drug use), likely country of transmission, Pre-Exposure Prophylaxis (PrEP) use, and the partner’s history of STIs. All information was collected anonymously and voluntarily. Blood samples were collected for HIV and syphilis testing. Urine, rectal, oropharyngeal and/or urogenital swabs were collected to test for CT and NG. The paper questionnaires and all biological samples were sent to Labor Krone, the national reference laboratory for syphilis, which was responsible for analysing all samples, digitising the paper questionnaires, and merging the data with test results using a unique ID number. The anonymised dataset, representing consultations rather than unique individuals, was submitted by the laboratory to the NRW Centre for Health (Landeszentrum Gesundheit NRW) every six months as an Excel line list.

### Data analysis

#### Determinations of STI positivity and the number of tests performed

HIV, CT and NG tests were considered performed if there was either a positive or negative test result available. Syphilis testing was considered performed if any of the test outcome categories were reported. HIV, CT and NG infections were considered positive if they were marked as positive in the dataset, regardless of the reason for testing; they were otherwise considered negative. Syphilis results were reported as a categorical variable: we considered syphilis infection only if the results indicated active syphilis infection (with, without, or unknown symptom status). Syphilis results categorised as sufficiently treated, latent syphilis or questionable treatment indication, follow-up for known syphilis, or false-positive screening, were not considered syphilis infections in this study.

#### STI test positivity

To estimate STI test positivity, the proportion of positive tests out of all tests performed was calculated for each infection. Percentages and 95% confidence intervals (CI) are reported for the study population overall and stratified by sex and key populations. Any man who reported MSM behaviour was classified as MSM, and all others were classified as heterosexual.

#### Populations of interest

Key populations (MSM, sex workers, and PWID) were identified based on STI transmission behaviours and high-risk exposures reported in the dataset. As individuals could report multiple exposures, the populations were not mutually exclusive.

#### Factors associated with STI test positivity

Data were analysed using RStudio software ^20^. We used univariable and multivariable logistic regression to examine factors associated with test positivity for each of HIV, syphilis, CT and NG. Variables with p-value <0.1 in univariable logistic regression analysis were included in the multivariable logistic regression model, using a forward stepwise selection process. Results were considered statistically significant in the final model if the p-value was less than 0.05.

### Ethics Approval

Ethical approval was provided by the London School of Hygiene & Tropical Medicine Research Ethics Committee (reference number: 27231). As these anonymous data are part of routine surveillance conducted by LZG.NRW, they are, by German law, exempt from needing ethical approval in Germany.

## Results

### Study population characteristics

Of the 11,560 consultations, the majority were attended by individuals who were German (72.8%), male (58.7%) and under 35 years old (72.1%). Most reported STI transmission behaviours were heterosexual (64.3%) or MSM (28.8%) contact, and transmission likely occurred in Germany (83.7%). Heterosexual exposure was the most frequently reported exposure category among both men and women, with the second most reported exposures being MSM contact in men (48.1%) and sex work in women (13.5%). PWID was reported as a transmission behaviour in fewer than 0.5% of consultations, and PrEP use was 5.3% in MSM. Information on STI transmission behaviour was missing for 700 (6.1%) consultations. The individuals’ characteristics recorded during consultations are shown in Table 1.

**Table 1:** Characteristics of individuals attending consultations at anonymous walk-in clinics in North-Rhine Westphalia state in 2021, by sex.

|  | Consultations<br>(N=11,560) | Women<br>(N=4,731) | Heterosexual<br>men<br>(N=3,517) | Men<br>MSM<br>(N=3,264) |
| --- | --- | --- | --- | --- |
| Sex<sup>1</sup> |  |  |  |  |
| Female | 4731 (40.9) | 4731 (100) | 0 (0) | 0 (0) |
| Male | 6781 (58.7) | 0 (0) | 3517 (100) | 3264 (100) |
| Other | 29 (0.3) | 0 (0) | 0 (0) | 0 (0) |
| Age group<br>(years)<sup>2</sup> |  |  |  |  |
| <25 | 2878 (24.9) | 1412 (29.8) | 711 (20.2) | 743 (22.8) |
| 25-34 | 5459 (47.2) | 2182 (46.1) | 1703 (48.4) | 1550 (47.5) |
| 35-49 | 2391 (20.7) | 895 (18.9) | 824 (23.4) | 661 (20.3) |
| 50-64 | 717 (6.2) | 216 (4.6) | 238 (6.8) | 263 (8.1) |
| 65+ | 78 (0.7) | 11 (0.2) | 31 (0.9) | 36 (1.1) |
| High-risk<br>exposure<sup>3</sup> |  |  |  |  |
| Heterosexual | 7432 (64.3) | 3971 (83.9) | 3020 (85.9) | 416 (12.7) |
| MSM<sup>7</sup> | 3330 (28.8) | 50 (1.1) | 0 (0) | 3264 (100) |
| FSF<sup>7</sup> | 284 (2.5) | 196 (4.1) | 39 (1.1) | 41 (1.3) |
| Sex work | 705 (6.1) | 639 (13.5) | 16 (0.5) | 44 (1.3) |
| Contact with<br>sex worker | 218 (1.9) | 21 (0.4) | 178 (5.1) | 19 (0.6) |
| PWID | 45 (0.4) | 11 (0.2) | 15 (0.4) | 19 (0.6) |
| Unknown | 700 (6.1) | 298 (6.3) | 394 (11.2) | 0 (0) |
| PrEP user<sup>4</sup> | 180 (1.6) | 4 (0.1) | 3 (0.1) | 173 (5.3) |
| Partner STI<br>status |  |  |  |  |
| Positive for one<br>or more STI | 770 (6.7) | 174 (3.7) | 214 (6.1) | 382 (11.7) |
| Transmission<br>location<sup>5</sup> |  |  |  |  |
| Germany | 9677 (83.7) | 3772 (79.7) | 2926 (83.2) | 2937 (90) |
| Country of<br>origin |  |  |  |  |
| Germany | 8413 (72.8) | 3248 (68.7) | 2684 (76.3) | 2454 (75.2) |
| Europe<sup>6</sup> | 1057 (9.1) | 696 (14.7) | 117 (3.3) | 239 (7.3) |
| Other Europe | 227 (2) | 107 (2.3) | 41 (1.2) | 77 (2.4) |
| Sub-Saharan<br>Africa | 220 (1.9) | 115 (2.4) | 80 (2.3) | 23 (0.7) |
| Middle East,<br>North Africa<br>and Turkey | 474 (4.1) | 109 (2.3) | 174 (4.9) | 189 (5.8) |
| Canada and<br>USA | 20 (0.2) | 5 (0.1) | 7 (0.2) | 8 (0.2) |
| East Asia &<br>Oceania | 124 (1.1) | 51 (1.1) | 23 (0.7) | 48 (1.5) |
| South Asia | 70 (0.6) | 9 (0.2) | 31 (0.9) | 30 (0.9) |
| Central &<br>South America | 172 (1.5) | 70 (1.5) | 23 (0.7) | 75 (2.3) |

MSM = men who have sex with men, FSF = females who have sex with females, PWID = people who inject drugs, EU/EEA = European Union/European Economic Area. PrEP = Preexposure prophylaxis
<sup>1</sup> Sex was determined based on the questionnaire – male, female or other (without further specification).
<sup>2</sup> Total proportions do not add to 100% due to missing data.
<sup>3</sup> Total proportions do not add to 100% because reported STI transmission behaviours were not mutually exclusive; multiple behaviours could be recorded for a single record.
<sup>4</sup> Uses PrEP regularly or occasionally.
<sup>5</sup> Location where the transmission likely took place.
<sup>6</sup> EU/EEA and UK, not including Germany.
<sup>7</sup> MSM and FSF were reported as transmission behaviours in women and men, respectively. In all records where FSF was reported for men or MSM was reported for women, heterosexual transmission behaviour was also documented.

### Number of STI tests performed

All four STIs were tested for during 45% of consultations, while only one STI was tested for in 14% of consultations. HIV was the most frequently tested infection (84%), followed by syphilis (69.5%), and CT/NG (65%). Table 2 summarises the number of tests performed by sex and key population group.

**Table 2:** Number of STI tests performed and test positivity by sex and high-risk exposure group.

|  |  | HIV | Syphilis | Chlamydia | Gonorrhoea | Any STI<sup>1</sup> | Coinfections<sup>1</sup> |
| --- | --- | --- | --- | --- | --- | --- | --- |
| All consultations (N=11,560) | n | 9716 | 8030 | 7522 | 7518 |  |  |
|  | % (95% CI) | 0.6 (0.5-0.8) | 0.9 (0.7-1.1) | 6.4 (5.8-6.9) | 2.7 (2.3-3.1) | 6.6 (6.1-7.0) | 0.4 (0.3-0.5) |
| Women (N=4,731) | n | 4031 | 3031 | 3072 | 3075 |  |  |
|  | % (95% CI) | 0.2 (0.1-0.4) | 0.3 (0.2-0.6) | 5.8 (5.0-6.6) | 1.5 (1.1-2.0) | 5.0 (4.4-5.6) | 0.1 (0-0.2) |
| Heterosexual sex in men (N=3,517) | n | 2967 | 2087 | 2179 | 2178 |  |  |
|  | % (95% CI) | 0.3 (0.2-0.6) | 0.4 (0.2-0.8) | 5.1 (4.2-6.1) | 0.6 (0.3-1.0) | 4.1 (3.5-4.9) | 0 (0-0.2) |
| MSM (N=3,264) | n | 2676 | 2873 | 2234 | 2228 |  |  |
|  | % (95% CI) | 1.5 (1.1-2.0) | 1.9 (1.4-2.4) | 8.3 (7.3-9.5) | 6.3 (5.4-7.4) | 11.6 (10.5-12.7) | 1.2 (0.9-1.7) |
| Sex work (N=705) | n | 546 | 556 | 554 | 554 |  |  |
|  | % (95% CI) | 0.9 (0.4-2.1) | 1.1 (0.5-2.3) | 9.2 (7.1-11.9) | 5.2 (3.7-7.4) | 12.6 (10.4-15.3) | 0.3 (0.1-1.0) |
| PWID (N=45) | n | 40 | 39 | 15 | 15 |  |  |
|  | % (95% CI) | 5 (1.4-16.5) | 2.6 (0.5-13.2) | 0 (0-20.4) | 0 (0-20.4) | 6.7 (2.3-17.9) | 0 (0-7.9) |
| Unknown exposure/risk (N=906) | n | 769 | 621 | 484 | 484 |  |  |
|  | % (95% CI) | 0.7 (0.3-1.7) | 0.7 (0.2-2.0) | 5.4 (3.4-8.5) | 2.9 (1.5-5.4) | 4.4 (3.1-6.2) | 0.3 (0.1-1.0) |

N = number of consultations, n = number of tests performed, % = test positivity, 95% CI = 95% Confidence intervals, MSM = men who have sex with men, PWID = people who inject drugs, STI = sexually transmitted infection
<sup>1</sup> The denominator for calculating the proportion of individuals diagnosed with any STI or with coinfections was the total number of consultations per group, regardless of the number of tests performed per consultation.

### STI test positivity

Overall, at least one STI was detected during 6.6% (95% CI 6.1-7.0%) of consultations. The highest test positivity was observed for CT (6.4%, 95% CI 5.8-6.9), followed by NG (2.7%, 95% CI 2.3-3.1). Syphilis and HIV were both detected in <1% of consultations (Table 2).

Among MSM and sex workers, CT was the most commonly detected infection (8.3%, 95% CI 7.3-9.5 and 9.2%, 95% CI 7.1-11.9, respectively). High test positivity was also observed for NG in MSM and sex workers (6.3%, 95% CI 5.4-7.4 and 5.2%, 95% CI 3.7- 7.4, respectively). In PWID, no CT/NG infections were detected, whereas test positivity for HIV and syphilis was 5.0% (95% CI 1.4-16.5) and 2.6% (95% CI 0.5-13.2), respectively. For individuals with unknown high-risk exposure, the test positivity for CT and NG respectively was 5.4% and 2.9%.

### Coinfections

Coinfections were detected in 47 (0.4%) consultations, of which 40 (85%) were in MSM (Figure 1). The most common simultaneously occurring infections were CT and NG (68%), which were reported in MSM and sex workers, as well as in individuals without information on high-risk exposure. The other co-infection combinations seen were all detected in MSM. Triple STI coinfections (CT-NG-syphilis and HIV-syphilis-NG) were detected at two consultations (4%).

**Figure 1:**
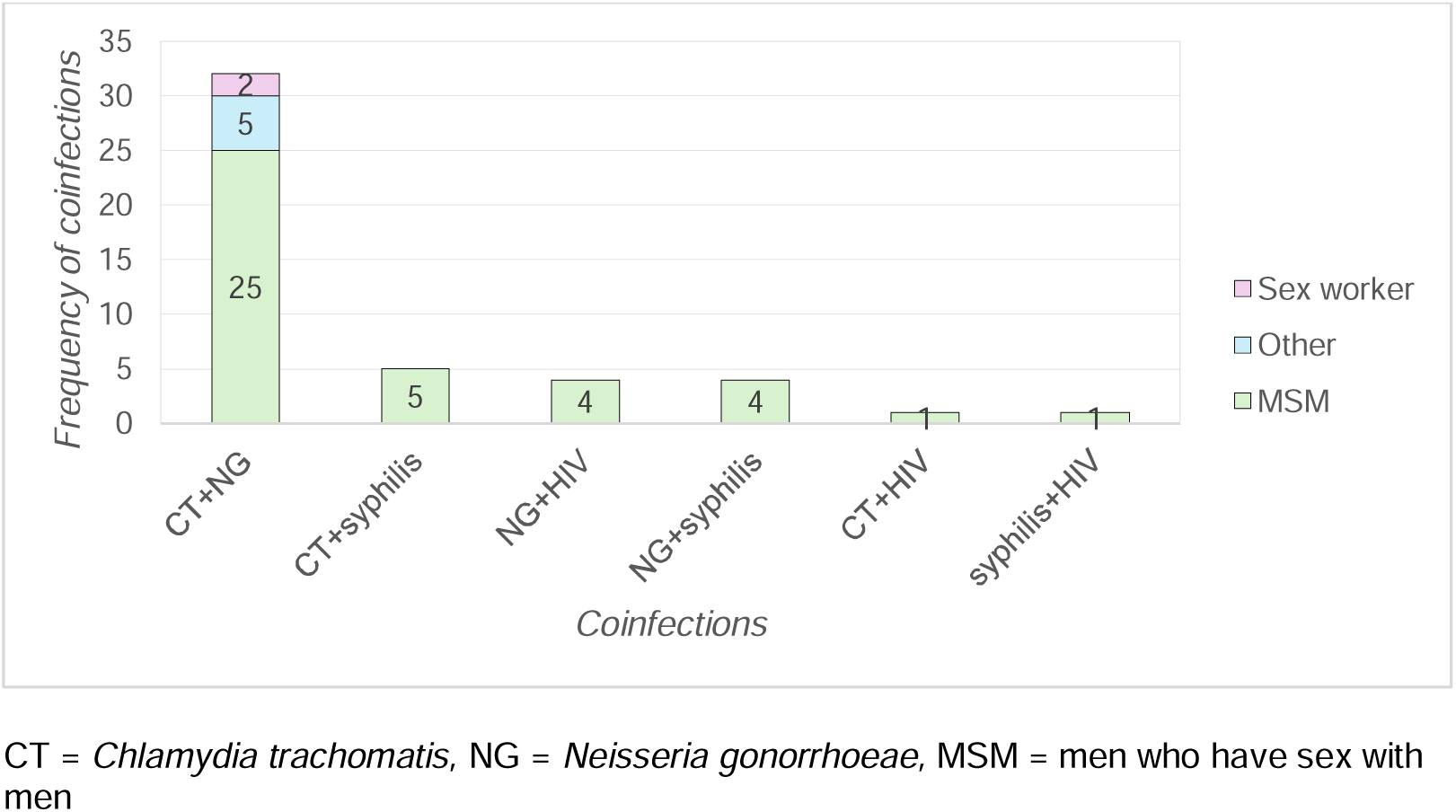
Frequency of STI coinfections by population group^1^ ^1^ No coinfections were diagnosed in people who inject drugs; category “Other” includes individuals who did not report MSM, sex work, or injection drug use as transmission behaviours, including individuals with unknown high-risk exposure.

### Factors associated with HIV test positivity

Factors with strong association with HIV test positivity were: being aged 35-49 years (aOR: 2.31, 95% CI 1.21-4.41) or 50-64 years (aOR: 2.88, 95% CI 1.17-6.46) compared to the reference 25-34 year age group; MSM transmission behaviour (aOR: 7.29, 95% CI 3.87-14.6); PWID (aOR: 9.11, 95% CI 1.41-33.1); partner positive for an STI (aOR: 2.16, 95% CI 1.0-4.26), and being from a Central Asian (aOR: 3.94%, 95% CI 1.15-10.3), Sub-Saharan African (aOR: 10.8, 95% CI 3.55-27.4) or Latin American (aOR: 3.47, 95% CI 0.81-10.1) country (Table 3).

**Table 3:** Factors associated with HIV, syphilis, chlamydia, gonorrhoea and any STI in multivariable logistic regression.

|  | HIV |  | Syphilis |  | Chlamydia |  | Gonorrhoea |  | Any STI |  |
| --- | --- | --- | --- | --- | --- | --- | --- | --- | --- | --- |
|  | aOR (95% CI) | p-value | aOR (95% CI) | p-value | aOR (95% CI) | p-value | aOR (95% CI) | p-value | aOR (95% CI) | p-value |
| Sex |  |  |  |  |  |  |  |  |  |  |
| Male |  |  | 7.02 (1.21, 133) | 0.072 |  |  |  |  | 1.05 (0.80, 1.36) | 0.7 |
| Female |  |  | Ref. | Ref. |  |  |  |  | Ref. | Ref. |
| Other |  |  | 0 (0.00, 27.1B) | >0.9 |  |  |  |  | 0 | >0.9 |
| Age group (years) |  |  |  |  |  |  |  |  |  |  |
| <25 | 0.54 (0.20, 1.35) | 0.2 |  |  | 1.12 (0.87, 1.44) | 0.4 |  |  |  |  |
| 25-34 | Ref. | Ref. |  |  | Ref. | Ref. |  |  |  |  |
| 35-49 | 2.31 (1.21, 4.41) | 0.011 |  |  | 0.72 (0.52, 0.98) | 0.042 |  |  |  |  |
| 50-64 | 2.88 (1.17, 6.46) | 0.014 |  |  | 0.77 (0.44, 1.27) | 0.3 |  |  |  |  |
| 65+ | 0 (0.00, >19.86M) | >0.9 |  |  | 0.51 (0.03, 2.49) | 0.5 |  |  |  |  |
| STI transmission behaviours<br>(Reference: STI behaviour not reported) |  |  |  |  |  |  |  |  |  |  |
| Heterosexual |  |  |  |  | 0.61 (0.49, 0.76) | <0.001 | 0.54 (0.30, 0.94) | 0.036 | 0.66 (0.49, 0.89) | 0.006 |
| MSM | 7.29 (3.87, 14.6) | <0.001 | 5.19 (2.26, 14.7) | <0.001 |  |  | 5.45 (2.93, 10.0) | <0.001 | 1.93 (1.38, 2.68) | <0.001 |
| FSF |  |  |  |  |  |  |  |  |  |  |
| Sex work |  |  |  |  | 1.86 (1.21, 2.76) | 0.003 | 4.22 (2.09, 8.01) | <0.001 | 2.77 (1.91, 3.96) | <0.001 |
| Contact with sex worker | 4.18 (0.66, 14.5) | 0.056 |  |  |  |  |  |  |  |  |
| PWID | 9.11 (1.41, 33.1) | 0.004 |  |  |  |  |  |  |  |  |
| PrEP Use |  |  |  |  |  |  |  |  |  |  |
| Yes |  |  |  |  |  |  | 2.7 (1.50, 4.61) | <0.001 |  |  |
| No |  |  |  | Ref. |  |  | Ref. | Ref. |  |  |
| Partner STI+ |  |  |  |  |  |  |  |  |  |  |
| Yes | 2.16 (1.00, 4.26) | 0.036 | 3.04 (1.54, 5.67) | <0.001 | 1.79 (1.26, 2.49) | <0.001 |  |  | 1.71 (1.30, 2.22) | <0.001 |
| No | Ref. | Ref. | Ref. | Ref. | Ref. | Ref. |  |  | Ref. |  |
| Likely transmission location |  |  |  |  |  |  |  |  |  |  |
| Germany |  |  |  |  |  |  |  |  |  |  |
| Abroad |  |  |  |  |  |  |  |  |  |  |
| Origin |  |  |  |  |  |  |  |  |  |  |
| Europe | Ref. | Ref. |  |  |  |  |  |  |  |  |
| Central Asia | 3.94 (1.15, 10.3) | 0.012 |  |  |  |  |  |  |  |  |
| Sub-Saharan Africa | 10.8 (3.55, 27.4) | <0.001 |  |  |  |  |  |  |  |  |
| MENA | 1.94 (0.57, 4.99) | 0.2 |  |  |  |  |  |  |  |  |
| North America | 0 | >0.9 |  |  |  |  |  |  |  |  |
| East Asia | 1.81 (0.10, 8.72) | 0.6 |  |  |  |  |  |  |  |  |
| South Asia | 0 (0.00, 1.82B) | >0.9 |  |  |  |  |  |  |  |  |
| Latin America | 3.47 (0.81, 10.1) | 0.045 |  |  |  |  |  |  |  |  |

MSM = men who have sex with men, FSF = females who have sex with females, PWID = people who inject drugs, EU/EEA = European Union/European Economic Area. PrEP = Preexposure prophylaxis, CI=Confidence intervals, OR = Odds Ratio, aOR=adjusted odds ratios, MENA = Middle East and North Africa

### Factors associated with acute syphilis test positivity

MSM transmission behaviours (aOR: 5.19, 95% CI 2.26-14.7) and partner positive for an STI (aOR: 3.04, 95% CI 1.54-5.67) had been strongly associated with acute syphilis test positivity (Table 3).

### Factors associated with CT test positivity

Factors independently associated with CT test positivity were sex work (1.86, 95% CI 1.21-2.76) and partner positive for an STI (aOR: 1.79, 95% CI 1.26-2.49). Being aged 35-49 years versus 25-34 years (aOR: 0.72, 95% CI 0.52-0.98), and heterosexual transmission behaviour (aOR: 0.61, 95% CI 0.49-0.76) were associated with lower odds of being CT-positive.

### Factors associated with NG test positivity

MSM (aOR: 5.45, 95% CI 2.93-10.0), sex work (aOR: 4.22, 95% CI 2.09-8.01) and PrEP use (aOR: 2.7, 95% CI 1.50-4.61) were independently associated with NG test positivity. Heterosexual transmission behaviours (aOR: 0.54, 95% CI 0.30-0.94) were associated with lower odds of being NG-positive.

## Discussion

Our analysis of sentinel surveillance data highlights a substantial burden of STIs among individuals tested at anonymous walk-in clinics in NRW, particularly among MSM and sex workers. Differences in STI test positivity across key population groups highlight the need for targeted prevention and testing strategies.

Overall, 6.6% of all consultations resulted in a diagnosis of at least one STI. The test positivity was the highest for CT (6.4%), followed by NG (2.7%). This pattern with CT as the most frequently detected STI is consistent with broader European surveillance data ^3^. The higher test positivity in our study likely reflects the high-risk of the population and the often asymptomatic nature of CT infections. The NG test positivity observed remains a concern, particularly given increasing antimicrobial resistance globally ^21^. While the value of CT/NG screening in asymptomatic individuals is debated ^22–24^, the known association with adverse reproductive and sexual health outcomes supports continued screening in high-risk populations ^25, 26^. Syphilis test positivity (0.9%) emphasises its ongoing relevance as a public health priority, aligning with global elimination strategies given the increasing global incidence ^27, 28^. Interestingly, syphilis test positivity among MSM was lower than some previous German estimates ^11, 15, 29, 30^, possibly reflecting differing case definitions and our focus on active infections rather than including previously treated cases. While this provides a more precise estimate of transmissible infections, it may underestimate the overall disease burden. HIV test positivity of 0.6% may be a reflection of effective prevention and treatment programmes. However, elevated rates in key populations indicate persistent disparities. Germany has made good progress toward achieving the UNAIDS 95-95-95 targets, achieving 99% treatment coverage and 96% viral suppression among diagnosed individuals, but only 92% of people living with HIV are diagnosed ^31^, highlighting gaps in case detection.

MSM were disproportionately affected by all STIs, with the highest positivity rates of CT (8.3%) and NG (6.3%). Previous research attributed high STI rates among MSM to dense sexual networks, condomless sex, and barriers to regular screening ^15, 16^. Although rare (0.4%), coinfections were predominantly observed among MSM. This is consistent with published research highlighting MSM as a key population ^13^, and may reflect overlapping high-risk sexual behaviours, including condomless anal intercourse, group sex, and chemsex practices ^13, 32^. Sex workers had a high STI burden, particularly for CT (9.2%) and NG (5.2%). Factors previously linked to high STI positivity in this group include structural vulnerabilities, substance abuse and limited access to healthcare ^33–35^. PWID, although the smallest subgroup (n=45), had the highest HIV (5%) and syphilis (2.6%) test positivity, suggesting unmet prevention needs. No CT/NG infections were detected in PWID, possibly reflecting a small sample size and limited testing, rather than a true absence of infection.

MSM behaviours, sex work, and having an STI-positive partner were associated with increased likelihood of testing STI-positive, although the strength and direction of association varied by infection. These findings demonstrate that STI prevention strategies tailored to both population and pathogen are needed, as one-size-fits-all approaches may overlook important behavioural and demographic differences, potentially undermining intervention effectiveness. We observed substantial variability in the range of STI tests conducted during consultations, suggesting inconsistent application of risk assessment protocols. For example, PWID were tested less frequently for CT/NG despite evidence of vulnerability due to overlapping sexual and injecting risks, as well as structural barriers such as stigma, marginalisation, and limited healthcare access ^36–38^. This selective rather than comprehensive testing may have contributed to both under- and over-testing. In addition, PrEP users typically receive comprehensive STI screening through specialised HIV practices rather than anonymous walk-in clinics, which could explain the low representation of PrEP users in our dataset.

Information on symptoms was unavailable, limiting our understanding of the criteria considered to offer testing, and the contribution of asymptomatic versus symptomatic infections to overall test positivity. Since STI testing of asymptomatic individuals is often deprioritised, this may represent missed opportunities for early detection and interruption of ongoing transmission. The recently agreed standardised criteria for determining STI testing eligibility in NRW’s public health services ^39^ should improve consistency and resource efficiency.

This study had several limitations. Our findings cannot be generalised beyond users of anonymous STI clinics in NRW. As we could not control which variables were collected, data quality issues inherent in routine surveillance may have affected completeness and accuracy ^40, 41^. Moreover, missing information on sexual behaviours, symptoms, and healthcare access (e.g. insurance coverage), plus incomplete information on high-risk behaviours (6%), limited our assessment of whether those most in need of services were reached. Reporting bias may also have affected disclosure, resulting in some individuals underreporting high-risk behaviours due to stigma, and others potentially overstating exposures to meet triage criteria. We lack information on how many individuals who sought consultation were not tested, limiting our ability to assess selection bias.

Data were collected during the COVID-19 pandemic, when STI services were scaled down and further triage measures were implemented ^42^. These pandemic-related service disruptions further limit the generalisability of our findings as they may have introduced selection bias or inflated test positivity estimates in our sample. HIV and syphilis testing at NRW anonymous walk-in clinics nearly halved during the pandemic (source: internal analysis of monthly reports from LHAs). Similar declines in testing during the COVID-19 pandemic were observed elsewhere in Europe ^43^, alongside reported decreases in sexual activity and the number of casual sexual partners ^44–46^. The extent to which the observed drop in testing in NRW can be attributed to reduced service availability, lower demand for testing, or changes related to sexual behaviour remains uncertain.

Our findings highlight the importance of targeted, evidence-based STI prevention and management strategies, as well as the added value of monitoring through sentinel surveillance systems. The substantial disparities observed across key populations demonstrate that MSM-focused STI testing strategies should remain a priority, while simultaneously expanding comprehensive services for other high-risk groups. Future research priorities should investigate behavioural and structural drivers of STI transmission among key populations, and examine how intersecting vulnerabilities – including gender, sexual orientation, and involvement in sex work – shape different STI risk profiles to develop inclusive and non-stigmatising services for all genders. Enhancing data collection through expanded consultation questionnaires incorporating standardised behavioural indicators (partner type, sexual practices, condom use, substance use), symptoms, healthcare access and social determinants of health, would enable more nuanced risk assessments and more informed tailored, population-specific interventions.

To our knowledge, this is the first analysis of test positivity for HIV, syphilis, CT and/or NG among users of anonymous walk-in clinics across NRW, with a focus on key populations beyond MSM. Given the limited availability of comparable studies in this setting, our findings address important knowledge gaps and contribute to a better understanding of STI epidemiology among key population groups, aligning with the objectives of Germany’s STI strategy ^19^.

## Data Availability

All data produced in the present study are available upon reasonable request to the authors.

## Acknowledgements

We would like to thank the local public health authorities in North Rhine-Westphalia and the clinical staff for their efforts in completing the questionnaires and collecting samples for STI testing. We are also grateful to Labor Krone for performing the laboratory analyses and consolidating the clinical test results with the questionnaire data.

## References

1. Sinka K. The global burden of sexually transmitted infections. Clin Dermatol 2024; 42: 110–118. 20231223. DOI: 10.1016/j.clindermatol.2023.12.002.

2. Zheng Y, Yu Q, Lin Y, et al. Global burden and trends of sexually transmitted infections from 1990 to 2019: an observational trend study. Lancet Infect Dis 2022; 22: 541–551. 2021/12/24. DOI: 10.1016/S1473-3099(21)00448-5.

3. European Centre for Disease Prevention and Control. A systematic review and meta-analysis of the prevalence of chlamydia, gonorrhoea, trichomoniasis and syphilis in Europe. 2024. Stockholm: ECDC.

4. Selb R, Bremer V, Jansen K, et al. Einführung einer Meldepflicht für N. gonorrhoeae mit verminderter Empfindlichkeit gegenüber Azithromycin, Cefixim oder Ceftriaxon. Epid Bull 2020: 6–12. DOI: 10.25646/6525.

5. Federal Ministry of Justice. Gesetz zur Verhütung und Bekämpfung von Infektionskrankheiten beim Menschen (Infektionsschutzgesetz - IfSG), https://www.gesetze-im-internet.de/ifsg/BJNR104510000.html (accessed 26 September 2024).

6. Robert Koch-Institut. Infektionsepidemiologisches Jahrbuch meldepflichtiger Krankheiten für 2022. 2024. Berlin: Robert Koch-Institut.

7. Robert Koch Institut. Umsetzung der neu eingeführten Meldepflichten nach §7 Abs. 3 IfSG: Meldung von Neisseria gonorrhoeae und Chlamydia trachomatis (Serotypen L1-L3), https://www.rki.de/DE/Themen/Infektionskrankheiten/Infektionskrankheiten-A-Z/G/Gonorrhoe/Meldepflicht_IfSG.html (2023, accessed 27 April 2025).

8. Jacob L, Bohler F, Kalder M, et al. Prevalence and treatment of sexually transmitted diseases in gynecological practices in Germany: A retrospective study with more than 1,000,000 patients. Int J Clin Pharmacol Ther 2018; 56: 212–216. DOI: 10.5414/CP203173.

9. Jacob L, Duse DA and Kostev K. Prevalence and treatment of sexually transmitted infections in men followed by urologists in Germany - a cross sectional study with 347,090 men. Ger Med Sci 2018; 16: Doc03. 20180813. DOI: 10.3205/000265.

10. Lallemand A, Bremer V, Jansen K, et al. Prevalence of Chlamydia trachomatis infection in women, heterosexual men and MSM visiting HIV counselling institutions in North Rhine-Westphalia, Germany - should Chlamydia testing be scaled up? BMC Infect Dis 2016; 16: 610. 20161026. DOI: 10.1186/s12879-016-1915-2.

11. Muller MC, Usadel S, Zimmermann S, et al. Closing Sexual Health Service Gaps With a New Service Model in Germany: Performance of an on-Site Integrated, Cross-Sectoral, Low Threshold Sexually Transmitted Infections/HIV Counseling and Treatment Service. Front Public Health 2022; 10: 793609. 20220425. DOI: 10.3389/fpubh.2022.793609.

12. Skaletz-Rorowski A, Potthoff A, Nambiar S, et al. Sexual behaviour, STI knowledge and Chlamydia trachomatis (CT) and Neisseria gonorrhoeae (NG) prevalence in an asymptomatic cohort in Ruhr-area, Germany: PreYoungGo study. J Eur Acad Dermatol Venereol 2021; 35: 241–246. 20201029. DOI: 10.1111/jdv.16913.

13. Jansen K, Steffen G, Potthoff A, et al. STI in times of PrEP: high prevalence of chlamydia, gonorrhea, and mycoplasma at different anatomic sites in men who have sex with men in Germany. BMC Infect Dis 2020; 20: 110. 20200207. DOI: 10.1186/s12879-020-4831-4.

14. Dudareva-Vizule S, Haar K, Sailer A, et al. Prevalence of pharyngeal and rectal Chlamydia trachomatis and Neisseria gonorrhoeae infections among men who have sex with men in Germany. Sex Transm Infect 2014; 90: 46–51. 20130806. DOI: 10.1136/sextrans-2012-050929.

15. Bremer V, Haar K, Gassowski M, et al. STI tests and proportion of positive tests in female sex workers attending local public health departments in Germany in 2010/11. BMC Public Health 2016; 16: 1175. 20161121. DOI: 10.1186/s12889-016-3847-6.

16. Marcus U, Kollan C, Gunsenheimer-Bartmeyer B, et al. HIV-Jahresbericht 2023. Epid Bull 2024; 40: 3–20. DOI: 10.25646/12856.

17. Jansen K and Bremer V. Syphilis in Deutschland in den Jahren 2020–2022 – Neuer Höchststand von Infektionen nach Rückgang während der COVID-19- Pandemie. Epid Bull 2024; 7: 3–24. DOI: 10.25646/11907.

18. Nitschke H, Ludwig-Diouf B, Knappik A, et al. Anonyme STD-Sprechstunde versus Pflichtuntersuchung fur Prostituierte--was ist effektiv in der STD-Pravention? Gesundheitswesen 2006; 68: 686–691. 2007/01/03. DOI: 10.1055/s-2006-927257.

19. Federal Ministry of Health; the Federal Ministry for Economic Cooperation and Development. Integrated Strategy for HIV, Hepatitis B and C and Other Sexually Transmitted Infections. 2016. Berlin: Federal Ministry of Health and the Federal Ministry for Economic Cooperation and Development.

20. RStudio Team. RStudio: Integrated Development for R. Boston, MA: RStudio, Inc., 2019.

21. Unemo M, Lahra MM, Escher M, et al. WHO global antimicrobial resistance surveillance for Neisseria gonorrhoeae 2017-18: a retrospective observational study. Lancet Microbe 2021; 2: 627–636. 20210902. DOI: 10.1016/S2666-5247(21)00171-3.

22. Williams E, Williamson DA and Hocking JS. Frequent screening for asymptomatic chlamydia and gonorrhoea infections in men who have sex with men: time to re-evaluate? Lancet Infect Dis 2023; 23: e558–e566. 20230726. DOI: 10.1016/S1473-3099(23)00356-0.

23. Dukers-Muijrers N, Evers YJ, Hoebe C, et al. Controversies and evidence on Chlamydia testing and treatment in asymptomatic women and men who have sex with men: a narrative review. BMC Infect Dis 2022; 22: 255. 20220314. DOI: 10.1186/s12879-022-07171-2.

24. Kenyon C. How actively should we screen for chlamydia and gonorrhoea in MSM and other high-ST-prevalence populations as we enter the era of increasingly untreatable infections? A viewpoint. J Med Microbiol 2019; 68: 132–135. 20181206. DOI: 10.1099/jmm.0.000889.

25. Cantor A, Dana T, Griffin JC, et al. Screening for Chlamydial and Gonococcal Infections: Updated Evidence Report and Systematic Review for the US Preventive Services Task Force. JAMA 2021; 326: 957–966. DOI: 10.1001/jama.2021.10577.

26. Kenyon C, Herrmann B, Hughes G, et al. Management of asymptomatic sexually transmitted infections in Europe: towards a differentiated, evidence-based approach. Lancet Reg Health Eur 2023; 34: 100743. 20231026. DOI: 10.1016/j.lanepe.2023.100743.

27. Yu W, You X and Luo W. Global, regional, and national burden of syphilis, 1990-2021 and predictions by Bayesian age-period-cohort analysis: a systematic analysis for the global burden of disease study 2021. Front Med (Lausanne) 2024; 11: 1448841. 20240815. DOI: 10.3389/fmed.2024.1448841.

28. Rowley J, Vander Hoorn S, Korenromp E, et al. Chlamydia, gonorrhoea, trichomoniasis and syphilis: global prevalence and incidence estimates, 2016. Bull World Health Organ 2019; 97: 548–562P. 20190606. DOI: 10.2471/BLT.18.228486.

29. Streeck H, Jansen K, Crowell TA, et al. HIV pre-exposure prophylaxis was associated with no impact on sexually transmitted infection prevalence in a high-prevalence population of predominantly men who have sex with men, Germany, 2018 to 2019. Euro Surveill 2022; 27. DOI: 10.2807/1560-7917.ES.2022.27.14.2100591.

30. Jansen K, Schmidt AJ, Drewes J, et al. Increased incidence of syphilis in men who have sex with men and risk management strategies, Germany, 2015. Euro Surveill 2016; 21. DOI: 10.2807/1560-7917.ES.2016.21.43.30382.

31. an der Heiden M, Marcus U, Kollan C, et al. Estimate of the number of new HIV infections in 2022 and 2023 and the total number of people living with HIV in Germany at the end of 2023 Epid Bull 2024; 28: 3–20. DOI: 10.25646/12212.

32. Hess KL, Crepaz N, Rose C, et al. Trends in Sexual Behavior Among Men Who have Sex with Men (MSM) in High-Income Countries, 1990-2013: A Systematic Review. AIDS Behav 2017; 21: 2811–2834. DOI: 10.1007/s10461-017-1799-1.

33. Sherman SG, Tomko C, White RH, et al. Structural and Environmental Influences Increase the Risk of Sexually Transmitted Infection in a Sample of Female Sex Workers. Sex Transm Dis 2021; 48: 648–653. DOI: 10.1097/OLQ.0000000000001400.

34. Park JN, Gaydos CA, White RH, et al. Incidence and Predictors of Chlamydia, Gonorrhea and Trichomonas Among a Prospective Cohort of Cisgender Female Sex Workers in Baltimore, Maryland. Sex Transm Dis 2019; 46: 788–794. DOI: 10.1097/OLQ.0000000000001085.

35. Tesfie TK, Yismaw GA, Yirsaw BG, et al. Prevalence and associated factors of HIV among female sex workers in Eastern and Southern Africa: Systematic review and meta-analysis. PLoS One 2024; 19: e0313868. 20241202. DOI: 10.1371/journal.pone.0313868.

36. Marks LR, Reno H, Liang SY, et al. Value of Packaged Testing for Sexually Transmitted Infections for Persons who Inject Drugs Hospitalized With Serious Injection-Related Infections. Open Forum Infect Dis 2021; 8 20211111. DOI: 10.1093/ofid/ofab489.

37. Steffen G, Krings A and Zimmermann R. Prevalence of hepatitis B and C, HIV, and syphilis among people who inject drugs in Germany. European Journal of Public Health 2022; 32. DOI: 10.1093/eurpub/ckac131.200.

38. Brookmeyer KA, Haderxhanaj LT, Hogben M, et al. Sexual risk behaviors and STDs among persons who inject drugs: A national study. Prev Med 2019; 126: 105779. 20190715. DOI: 10.1016/j.ypmed.2019.105779.

39. Grotegut P, Spiekermann K, Wentzky S, et al. Etablierung von Kriterien für die in NRW landesfinanzierten Testungen auf sexuell übertragbaren Infektionen im öffentlichen Gesundheitsdienst. Das Gesundheitswesen 2025; 87: S32–S33. DOI: 10.1055/s-0045-1801955.

40. Newman LM, Samuel MC, Stenger MR, et al. Practical Considerations for Matching STD and HIV Surveillance Data with Data from Other Sources. Public Health Rep 2009; 124: 7–17. DOI: 10.1177/00333549091240S203.

41. Hemkens LG, Contopoulos-Ioannidis DG and Ioannidis JPA. Routinely collected data and comparative effectiveness evidence: promises and limitations. CMAJ 2016; 188: 158–164. 20160216. DOI: 10.1503/cmaj.150653.

42. van Bremen K, Monin M, Schlabe S, et al. Impact of COVID-19 on HIV late diagnosis in a specialized German centre. HIV Med 2022; 23: 1209–1213. 20221020. DOI: 10.1111/hiv.13426.

43. Simoes D, Stengaard AR, Combs L, et al. Impact of the COVID-19 pandemic on testing services for HIV, viral hepatitis and sexually transmitted infections in the WHO European Region, March to August 2020. Euro Surveill 2020; 25. DOI: 10.2807/1560-7917.ES.2020.25.47.2001943.

44. Masoudi M, Maasoumi R and Bragazzi NL. Effects of the COVID-19 pandemic on sexual functioning and activity: a systematic review and meta-analysis. BMC Public Health 2022; 22: 189. 20220128. DOI: 10.1186/s12889-021-12390-4.

45. van Bilsen WPH, Zimmermann HML, Boyd A, et al. Sexual Behavior and Its Determinants During COVID-19 Restrictions Among Men Who Have Sex With Men in Amsterdam. J Acquir Immune Defic Syndr 2021; 86: 288–296. DOI: 10.1097/QAI.0000000000002581.

46. Mourikis I, Kokka I, Koumantarou-Malisiova E, et al. Exploring the adult sexual wellbeing and behavior during the COVID-19 pandemic. A systematic review and meta-analysis. Front Psychiatry 2022; 13: 949077. 20220818. DOI: 10.3389/fpsyt.2022.949077.

